# Links between air pollution and COVID-19 in England

**DOI:** 10.1101/2020.04.16.20067405

**Authors:** Marco Travaglio, Yizhou Yu, Rebeka Popovic, Liza Selley, Nuno Santos Leal, Luis Miguel Martins

## Abstract

In December 2019, a novel disease, coronavirus disease 19 (COVID-19), emerged in Wuhan, People’s Republic of China. COVID-19 is caused by a novel coronavirus (SARS-CoV-2) presumed to have jumped species from another mammal to humans. This virus has caused a rapidly spreading global pandemic. To date, thousands of cases of COVID-19 have been reported in England, and over 25,000 patients have died. While progress has been achieved in managing this disease, the factors in addition to age that affect the severity and mortality of COVID-19 have not been clearly identified. Recent studies of COVID-19 in several countries identified links between air pollution and death rates. Here, we explored potential links between major air pollutants related to fossil fuels and SARS-CoV-2 mortality in England. We compared current SARS-CoV-2 cases and deaths recorded in public databases to both regional and subregional air pollution data monitored at multiple sites across England. We show that the levels of multiple markers of poor air quality, including nitrogen oxides and sulphur dioxide, are associated with increased numbers of COVID-19-related deaths across England, after adjusting for population density. We expanded our analysis using individual-level data from the UK Biobank and showed that particulate matter contributes to increased infectivity. We also analysed the relative contributions of individual fossil fuel sources on key air pollutant levels. The levels of some air pollutants are linked to COVID-19 cases and adverse outcomes. This study provides a useful framework to guide health policies in countries affected by this pandemic.

## INTRODUCTION

In December 2019, a high number of pneumonia cases with an unknown aetiology were detected in Wuhan, China. A molecular analysis of samples from affected patients revealed that their symptoms were caused by an infection with a novel coronavirus, later named severe acute respiratory syndrome (SARS) coronavirus (CoV) 2 (SARS-CoV-2), the pathogenic agent of coronavirus disease 19 (COVID-19) ^1^. Coronaviruses are a genus of enveloped, non-segmented, positive-sense RNA viruses belonging to the family Coronaviridae and classified within the Nidovirales order ^2^. Historically, illnesses caused by coronaviruses have ranged in severity, with some, including human coronaviruses-229E and -OC43, causing common cold symptoms, but SARS-CoV and Middle East respiratory syndrome coronavirus have initiated outbreaks of life-threatening pneumonia. While the initial symptoms of COVID-19 include fever with or without respiratory syndrome, a crescendo of pulmonary abnormalities may subsequently develop in patients ^3^. According to recent studies, most patients present with only a mild illness, but approximately 25% of hospital-admitted patients require intensive care because of viral pneumonia with respiratory complications ^4^.

While extensive research into the pathogenesis of COVID-19 suggests that the severe disease likely stems from an excessive inflammatory response ^5^, the exact predisposing factors contributing to an increased clinical severity and death in patients remain unclear. Individuals over the age of 60 years or with underlying health conditions, including cardiovascular and chronic respiratory diseases, diabetes, and cancer, are at the highest risk of a severe disease and death ^6^. Long-term exposure to air pollutants has been shown to be a risk factor mediating the pathogenesis of these health conditions ^7^. In fact, prolonged exposure to common road transport pollutants, including nitrogen oxides, sulphur dioxide and ground-level ozone, significantly exacerbates cardiovascular morbidity, diabetes, airway oxidative stress and asthma ^8,9^. These pollutants also cause a persistent inflammatory response and increase the risk of infection with viruses that target the respiratory tract ^10-12^. In addition, airborne particulate matter (PM) was recently shown to increase the viability of SARS-CoV-2, suggesting direct microbial pathogenic transmission through the air and increased opportunity for infection in highly polluted areas ^13^. The geographical patterns of COVID-19 transmission and mortality among countries, and even among regions of single countries, closely align with local levels of air pollutants ^11^. For example, increased contagiousness and COVID-19-related mortality in northern Italian regions, including Lombardia, Veneto and Emilia Romagna, have been correlated with high levels of air pollutants in these regions ^11^.

Here, we explored the relationship between air pollution and COVID-19 using an approach that combines both population- and individual-level data. We first investigated potential links between regional and subregional variations in air pollution and COVID-19-related deaths and cases in England by employing coarse and fine resolution methods. Next, we addressed the associations between several air pollutants and the risk of COVID-19 infection at the individual scale by analysing UK Biobank data obtained from a cohort of 1450 subjects. Finally, we modelled the relationship between several fossil-fuel burning sources and annualised daily measurements for multiple air pollutants to identify the major sources of air pollutants contributing to increased deaths in England.

## METHODS

### Data sources for COVID-19 deaths and cases

The number of patients infected with SARS-CoV-2 in England was obtained from Public Health England (PHE) and analysed according to the following statistical regions: London, Midlands, Northwest, Northeast and Yorkshire, Southeast, East, and Southwest England. Region-level data on the cumulative number of SARS-CoV-2-related deaths in England was retrieved from the National Health Service (NHS) (Table 1). This source provides one of the most comprehensive region-specific records of COVID-19-related deaths in England. The daily death summary included the number of deaths of patients who died in hospitals in England and had tested positive for SARS-CoV-2 at the time of death. While this online repository is updated daily, figures are subject to change due to a *post-mortem* confirmation of the diagnosis. Local authority-level data on the cumulative number of COVID-19 deaths in England was provided by the Office for National Statistics (ONS) (Table 1). This repository includes deaths of patients who died in care homes or other places outside hospitals. All deaths are recorded as the date of death rather than the day on which the death was announced. The cumulative number of local authority COVID-19 cases was provided by PHE (Table 1). Local authority-level data included the numbers of deaths and cases in England up to and including the 10th of April, approximately two weeks after the UK was placed into lockdown.

**Table 1.**
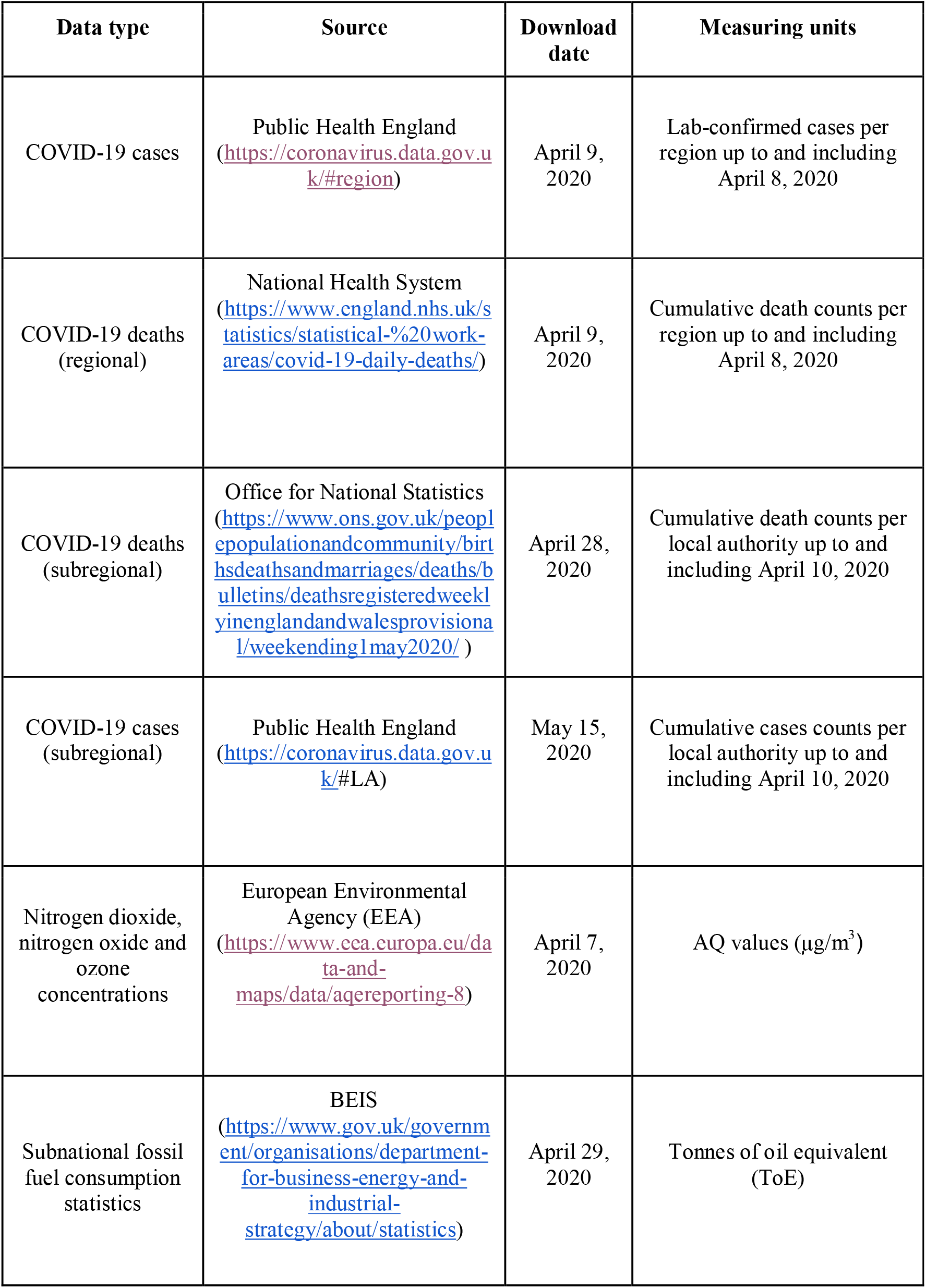
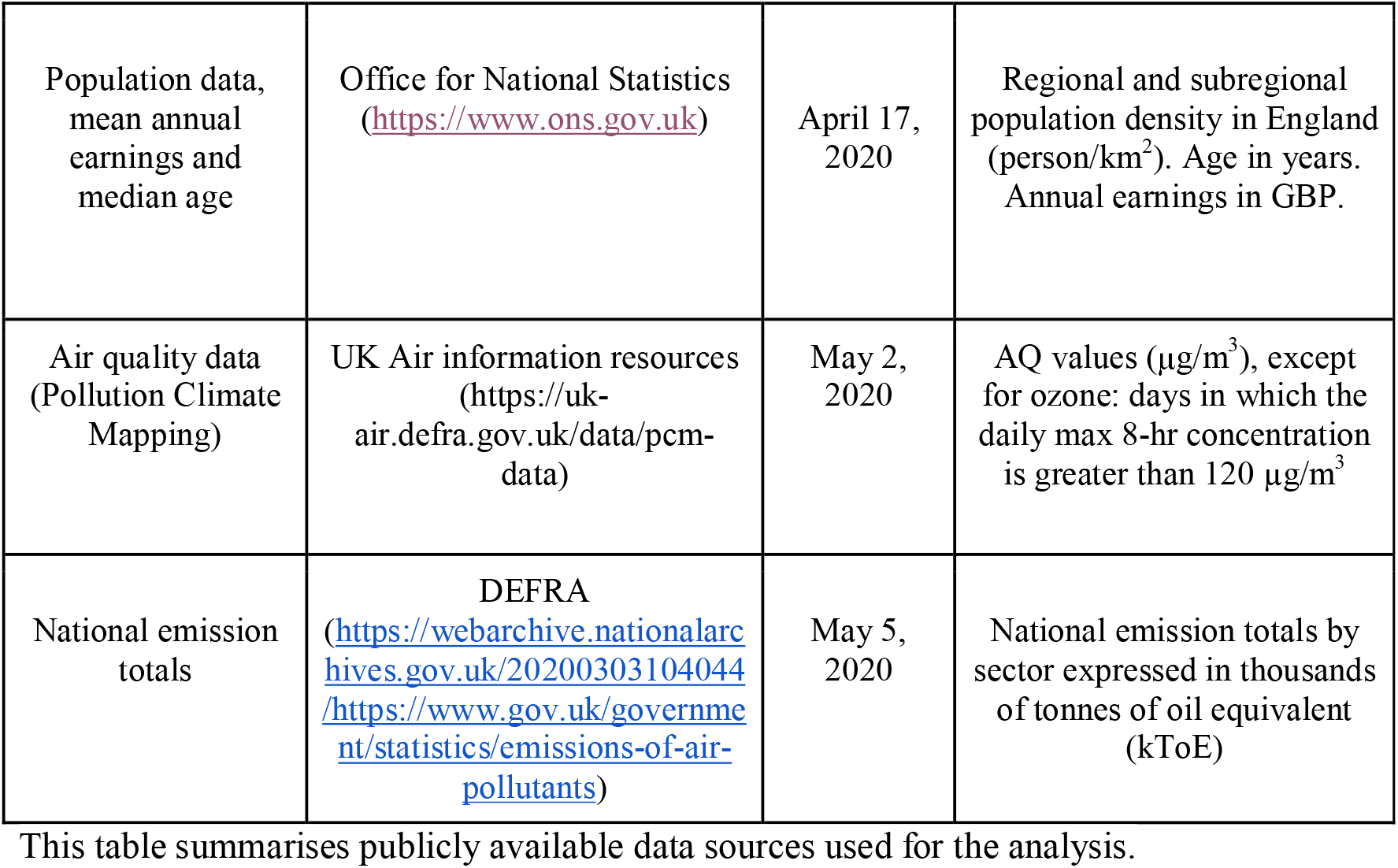
Summary of data sources.

### Data sources for air pollution levels

Air pollution data for 2018 were obtained from two sources. For the initial region-level analysis, we collected annual aggregated air quality (AQ) values determined by the European Environmental Agency based on direct observations obtained from multiple monitoring stations located across England. Due to incomplete or obsolete observations for several pollutants, we restricted our analysis to individual pollution indices for three major air pollutants, namely, nitrogen dioxide, nitrogen oxide and ozone, across the prespecified regions (Figure 2). Nitrogen dioxide, nitrogen oxide and ozone AQ values are reported in μg/m^3^ and represent the annual average of daily measurements for each air pollutant from 2018 to 2019 in each specified region. The identification of each monitoring station was matched to each available city by accessing the Department for Environment, Food and Rural Affairs (DEFRA) website (Figure 1). This website contains a resource called the DEFRA’s Air Quality Spatial Object Register, which allows users to view and retrieve information on the air quality-related spatial and non-spatial data objects from the UK’s Air Quality e-Reporting data holdings. The annual average values of daily measurements for each pollutant in each monitoring area were analysed to determine the effects of toxin exposure on the number of SARS-CoV-2 cases and deaths across England (Figure 1).

**Figure 1.**
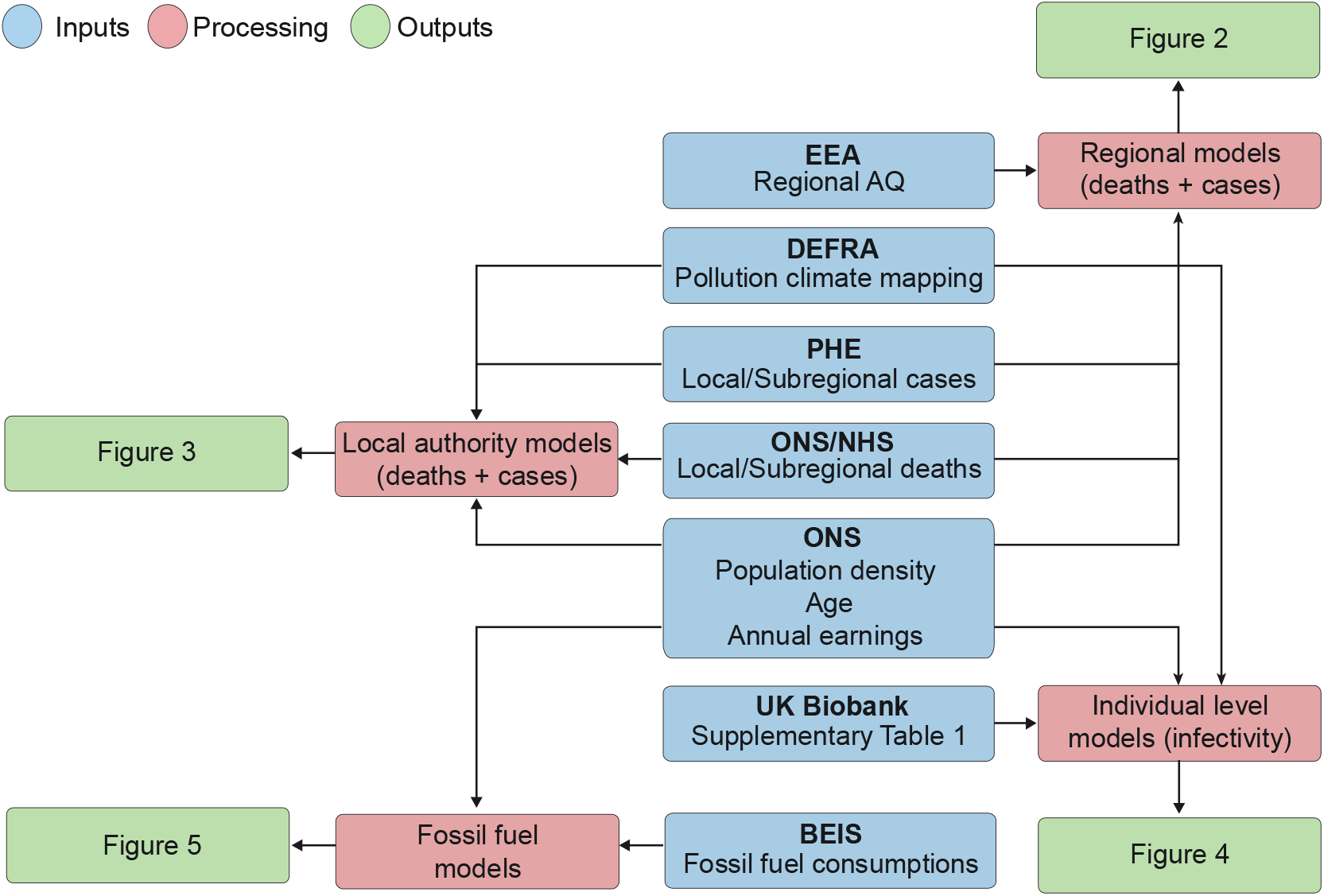
Analysis workflow. This flowchart summarizes how raw data were extrapolated, processed and analysed. Blue indicates data sources, whereas red and green indicate the type of model employed and the final output, respectively. Population density data (person/km^2^) were derived from ONS and used to account for region-specific differences in population size across England; COVID-19 case and death data were obtained from PHE, NHS and ONS, respectively. Air pollution data from each monitoring station were manually curated using DEFRA’s Air Quality Spatial Object Register and aggregated into statistical regions. ONS, Office for National Statistics; PHE, Public Health England; NHS, National Health Service; EEA, European Environmental Agency.

For the analysis at the level of local authorities, we used the Pollution Climate Mapping data from the UK Air Information Resources (Table 1). This repository contains information from hundreds of air quality stations located across England for multiple pollutant molecules (ozone, nitrogen oxides, PM_25_, PM_10_, and sulphur dioxide). All data represent annual average values of daily measurements for 2018 and are reported in μg/m^3^, except for ozone, whose metric is the number of days on which the daily max 8-hr concentration is greater than 120 μg/m^3^. A detailed quality report regarding this data is available at the following website: https://uk-air.defra.gov.uk/assets/documents/reports/cat09/1903201606 AQ0650 2017 MAAQ techni cal report.pdf. We obtained the longitude and latitude of each local authority using OpenCage Geocoder (https://opencagedata.com/). The air pollutant levels for each authority was approximated by averaging 10 values nearest the centre of authority. This area covers approximately 12 km^2^. Detailed descriptions of the methodology and analysis workflow are available in our GitHub repository. For the UK Biobank data, we matched the location coordinate each participant reported to their nearest modelled value. The level of each pollutant was measured less than 2 km away from the self-reported address.

### Subnational fossil fuel consumption data

Subnational fossil fuel consumption statistics were derived from the Department for Business, Energy and Industrial Strategy (BEIS) online data repository, which represents the most authoritative and up-to-date source of fossil fuel emissions in the UK. Local authority-level fossil fuel consumption estimates are produced as part of the National Atmospheric Emissions Inventory (NAEI) work programme on fuel consumption from road transport, manufacturing industry and other sources. Fuel consumption data from road transport is determined using a bottom-up approach that combines fleet-weighted fuel consumption factors (in g of fuel/km) for each main vehicle type (bus, cars, motorcycles, light-goods vehicles (LGV) and heavy-goods vehicles (HGV)) with traffic activity data provided by the Department for Transport (DfT). Estimates of road transport consumption are based on five vehicle types (buses, cars, motorcycles, HGV and LGV) and two fuel types (petrol and diesel). Road transport consumption is further categorised according to road class (motorways, A-roads and minor roads) to account for road-type variations in traffic volumes across the country.

Residual fuel consumption by the consuming sector was calculated by Ricardo Energy & Environment using the NAEI distribution maps and energy consumption estimates for point sources at known locations (https://assets.publishing.service.gov.uk/government/uploads/system/uploads/attachment_data/file/833214/UK_sub-national_residual_fuel_consumption_for_2005-2017_Estimates_of_non-gas_non-electricity_and_non-road_transport_energy.pdf). Residual fuels are defined as non-gas, non-electric and non-road transport fuels not used for the generation of electricity or road transport. This dataset is derived from the results of the NAEI and Greenhouse Gas Inventory (GHGI) survey conducted by Ricardo Energy & Environment on behalf of BEIS and excludes fuel used in aviation, shipping and power stations. Sources of fuel for this category included petroleum, coal, bioenergy and waste and the sectors considered included public administration, agriculture, industry, commercial, domestic and rail. Data for subnational fuel consumption statistics is reported in tonnes of oil equivalent (ToE), which is a unit of energy defined as the amount of energy released by burning one tonne of crude oil.

Annualised and weather-corrected gas consumption data were obtained from Xoserve, which generates annualised consumption estimates for all meter point reference numbers (MPRN) or gas meters on behalf of BEIS. The classification of domestic and non-domestic (commercial and industrial) is based on the gas industry standard cut-off point of 73,200 kWh. The weather correction factor used by Xoserve accounts for variations in regional temperature, domestic use and wind speed, enabling comparisons of gas use over time while controlling for changes in weather. Average domestic and industrial and commercial consumption are reported as sales per meter in kilowatt hours (kWh). Local authority-level gas statistics were obtained based on the aggregation of MPRN readings throughout England, generated as part of BEIS’s annual meter point gas data exercise.

### UK Biobank data sources

We used data from the UK Biobank under application #60124. Details regarding the geographical regions, recruitment processes, and other characteristics have been previously described ^14^, and are found on ukbiobank.co.uk. The UK Biobank has received ethical approval from the North West – Haydock Research Ethics Committee, 11/NW/0382 to gather data from participants. A detailed list of the variables analysed in the present study is presented in Supplementary Table 1. Notably, we defined hypertension using the criteria of a diastolic blood pressure ≥ 90 mmHg OR systolic blood pressure ≥ 140 mmHg.

### Regional heatmaps

Heatmap legends were generated using GraphPad Prism 8 (www.graphpad.com), and regions are labelled with the mapped colour values.

### Statistical analysis

In our regional exploratory analysis, we fitted generalised linear models to our data using COVID-19 deaths and cases as the outcomes and nitrogen oxide, nitrogen dioxide and ozone as the exposures of interest, adding the corresponding population density values as a confounding variable. Population density (person/km^2^) data correspond to subnational mid-year population estimates of the resident population in England for 2018 and excludes visitors or short-term immigrants (< 12 months). We modelled both the number of cases and deaths using negative binomial regression analyses since the response variables are overdispersed count data. We used the same model type for our subregional analysis, adding mean annual earnings and median age as confounding factors.

For the UK Biobank models, we fitted a binomial regression model because the response variable, COVID-positive or -negative, is defined as either 0 or 1.

For the analysis of fossil fuel consumption data, we employed multiple single pollutant models in which the dependent variables were the annual mean values of daily measurements of 4 air pollutants (nitrogen dioxide, nitrogen oxides, sulphur dioxide and ozone), and the independent variables included 21 sources of emissions from road transport, 9 from commercial and industrial sites and 2 from gas consumption. As these variables represent distinct groups, we computed the principal components of each group to explore the potential contribution of each group to the increased concentration of individual air pollutants as previously described ^15,16^. We used an iterative variable selection procedure combining unsupervised stepwise forward and stepwise backward regression analyses to further determine the individual contribution of each pollution source. Stepwise regression is commonly used in air pollution studies^17^ and was therefore used to select the most suitable predictor or combination of predictors within each polluting category. Generalized linear models of the gamma family were utilized for positively skewed, non-negative continuous response variables (nitrogen dioxide, nitrogen oxide and sulphur dioxide) using the log link function. A generalized linear model of the Gaussian family was applied to ozone data. Methods for assessing the fit of the model included residual analyses, diagnostic tests, and information criterion fit statistics. The goodness of fit of each regression model was determined using the log-likelihood and Akaike Information Criterion (AIC) statistics.

For all models, we calculated the odds or risk ratios and their 95% confidence intervals to quantify the effects of the independent variables on the response variables. The models were built using the MASS package (www.stats.ox.ac.uk/pub/MASS4/) in R. The comparison tables were generated using the Stargazer package ^18^. The analysis source code, detailed quality checks as well as all Supplementary material is available in GitHub (https://github.com/M1gus/AirPollutionCOVID19). The analysis notebook is available on the following link: https://m1gus.github.io/AirPollutionCOVID19/. Statistical significance was defined as *p* ≤ 0.05.

## RESULTS

### A link between regional nitrogen oxide and ozone levels and COVID-19 in England

We analysed the associations between cumulative numbers of COVID-19 cases and deaths with the concentrations of three major air pollutants recorded between 2018 and 2019, when no COVID-19 cases were reported. Due to differences in data availability for each air pollutant, we only included annual mean values of daily measurements, which was the most consistent aggregation type reported for all air pollutants described in this analysis. We started by analysing publicly available data from seven regions in England (Table 1), where a minimum of 2,000 SARS-CoV-2 infections and 200 deaths were reported by PHE from February 1 to April 8, 2020, approximately two weeks after the UK was placed into lockdown (Figure 1).

The spatial pattern of COVID-19 deaths matched the geographical distribution of COVID-19-related cases, with the largest numbers of COVID-19 deaths occurring in London and in the Midlands (Figure 2). According to previous studies, those two areas present the highest annual average concentration (μg/m^3^) of nitrogen oxides ^19^. In addition, ground-level ozone concentrations have been previously shown to vary significantly with latitude and altitude, depending on the concentration of ozone in the free troposphere, long-range transport and emission of its precursor ^20^. Therefore, we sought to determine if spatial variations in the levels of nitrogen oxides, in particular nitrogen dioxide (NO_2_) and nitrogen oxide (NO), as well as ground-level ozone concentrations in England are associated with increased numbers of COVID-19 infections and mortality. We applied a negative binomial regression model to estimate the association between each air pollutant with the cumulative number of both COVID-19 cases and deaths at the regional level (Supplementary Tables 2 and 3). The model was chosen based on the data type (count data) and log likelihood and AIC scores ^21^. Population density, a confounding factor, was added to this model as an independent variable to account for differences in the number of inhabitants across regions. The levels of nitrogen oxide and nitrogen dioxide are significant predictors of COVID-19 cases (*p* < 0.05), independent of the population density (Supplementary Table 2). We next applied a similar method to assess the association with the number of COVID-19 deaths (Supplementary Table 3). Ozone, nitrogen oxide and nitrogen dioxide levels are significantly associated with COVID-19 deaths, together with the population density.

**Figure 2.**
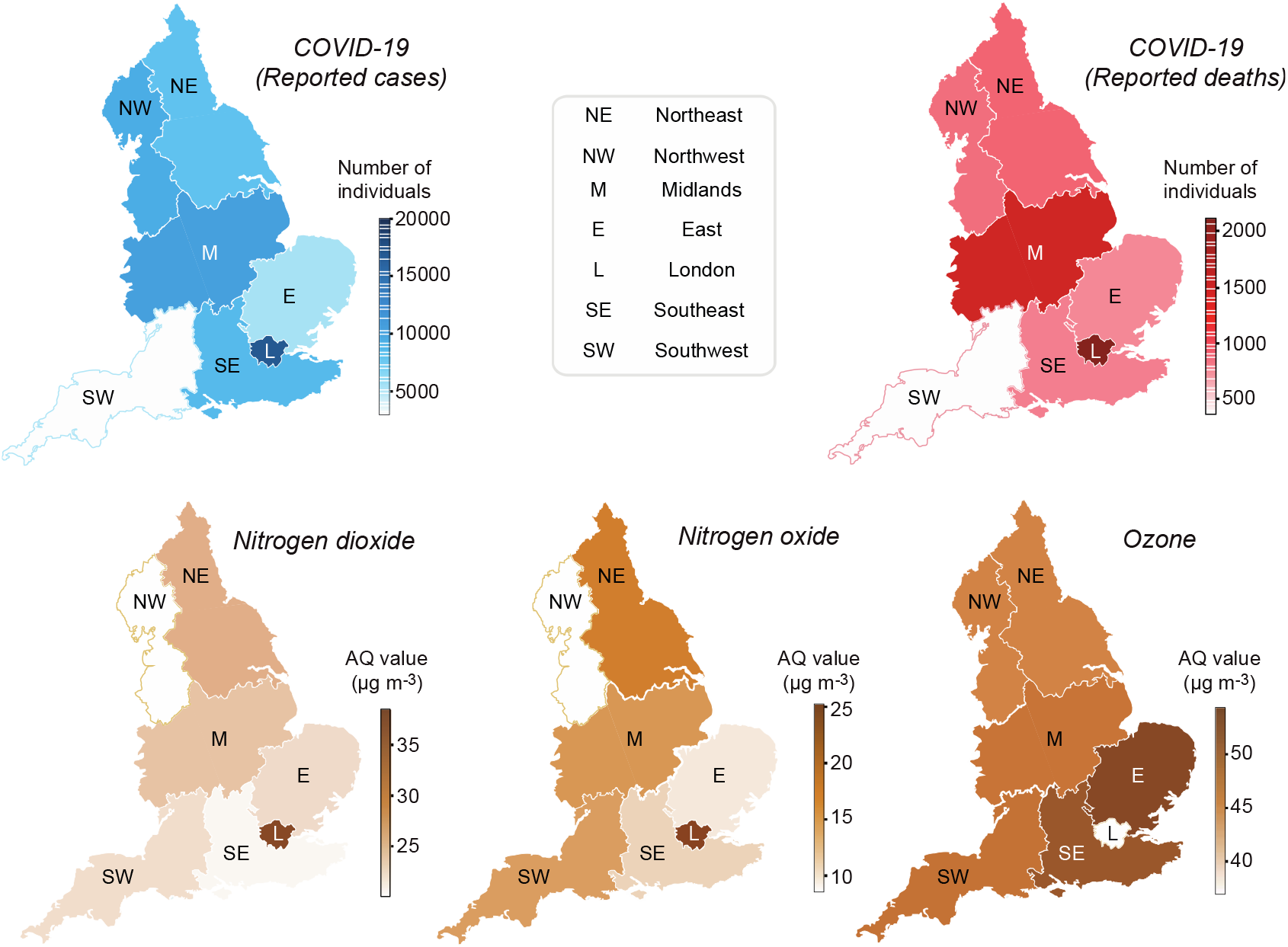
Regional heatmaps of COVID-19 and pollutants. Regional English heatmaps of reported deaths and diagnosed COVID-19 cases through April 8, 2020 (top row), as well as AQ values for the indicated pollutants (bottom row).

Taken together, the negative binomial regression models of both COVID-19 cases and deaths (Supplementary Tables 2 and 3) show that nitrogen dioxide, nitrogen oxide and ozone levels are significant predictors of COVID-19-related death, after accounting for the population density. This study provides the first evidence that SARS-CoV-2 cases and deaths are associated with regional variations in air pollution across England.

### Sulphur dioxide is a main contributor to increased numbers of COVID-19 deaths and cases at the subregional level

We next sought to increase both the resolution and accuracy of our analysis. We gathered data on COVID-related cases and deaths from all the local authorities in England and expanded the number of the pollutant species (n=6). We also retrieved the longitude and latitude for each local authority. The levels of ozone, nitrogen oxide, nitrogen dioxide, PM with aerodynamic diameters of 2.5 and 10 pm (PM_2_._5_ and PM_10_, respectively), and sulphur dioxide are reported as averages of the 10 values measured nearest the centre of each local authority in England. Local authority-level population density, mean annual earnings and age in 2018 were included as potential confounding variables (Figure 1). We calculated the estimated regression coefficients of each variable and their respective mortality and infectivity rate ratios (Figure 3 and Supplementary Tables 4 and 5) relative to the different air pollutants mentioned. Higher nitrogen or sulphur dioxide levels predict an increase in COVID-19 deaths and cases. The levels of sulphur dioxide have a mortality rate ratio of 1.172 [95% confidence interval (CI): 1.005-1.369] and infectivity rate ratio of 1.316 [95% CI: 1.141 - 1.521], indicating that a 1 μg/m^3^ increase in the sulphur dioxide concentration will lead to 17.2% more deaths and 31.6% more cases. Both the levels of nitrogen oxides and dioxide show mortality and infectivity rate ratios of approximately 1.03 (Figure 3). The incidence rate ratios of cases and deaths for ozone levels are less than 1, indicating that higher ozone levels lead to lower numbers of deaths and cases. PM_2.5_ and PM_10_ are negatively associated with the number of cases, and they are not significant predictors of the number of COVID-19-related deaths.

**Figure 3.**
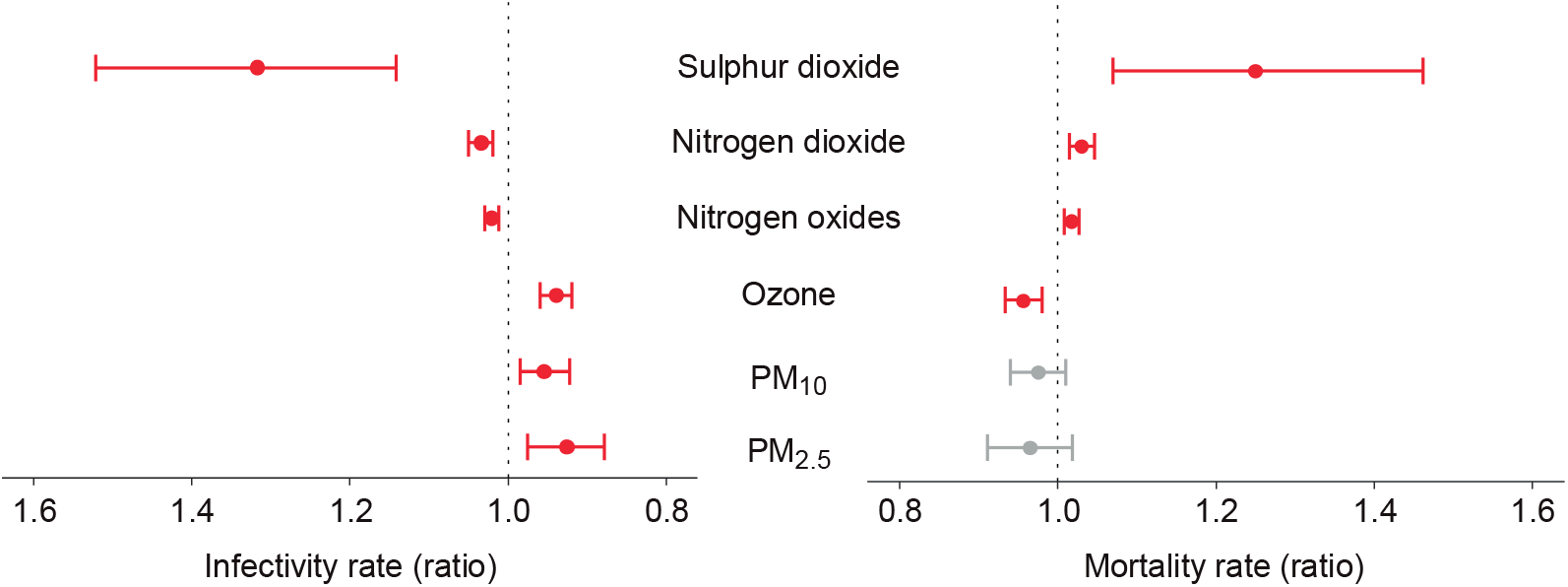
Cases and deaths in local authorities. Summary of infectivity and mortality rate ratios and respective 95% CIs at the local authority level. Red indicates significant associations (*p* ≤ 0.05), while grey lines show a lack of significance (*p* > 0.05). See also Supplementary Table 4 for a detailed description of the model.

### Levels of PM pollutants and nitrogen oxides are associated with an increase in SARS-CoV2 infections in UK Biobank participants living in England

We next used information from the UK Biobank to further assess whether people exposed to increased pollution levels are more likely to contract SARS-CoV-2 at the individual scale. This resource contains data from more than half a million UK volunteers recorded across multiple years. As of the writing of this paper, the UK Biobank dataset contains COVID-19 tests for 1,450 participants, of whom 669 were diagnosed as positive for COVID-19. The location of each subject included in the analysis is shown in Figure 4A. Compared to the local authority case model, the UK Biobank analysis provides a higher resolution air pollution estimate (less than 2 km away from their self-reported address) and includes potentially asymptomatic cases.

**Figure 4.**
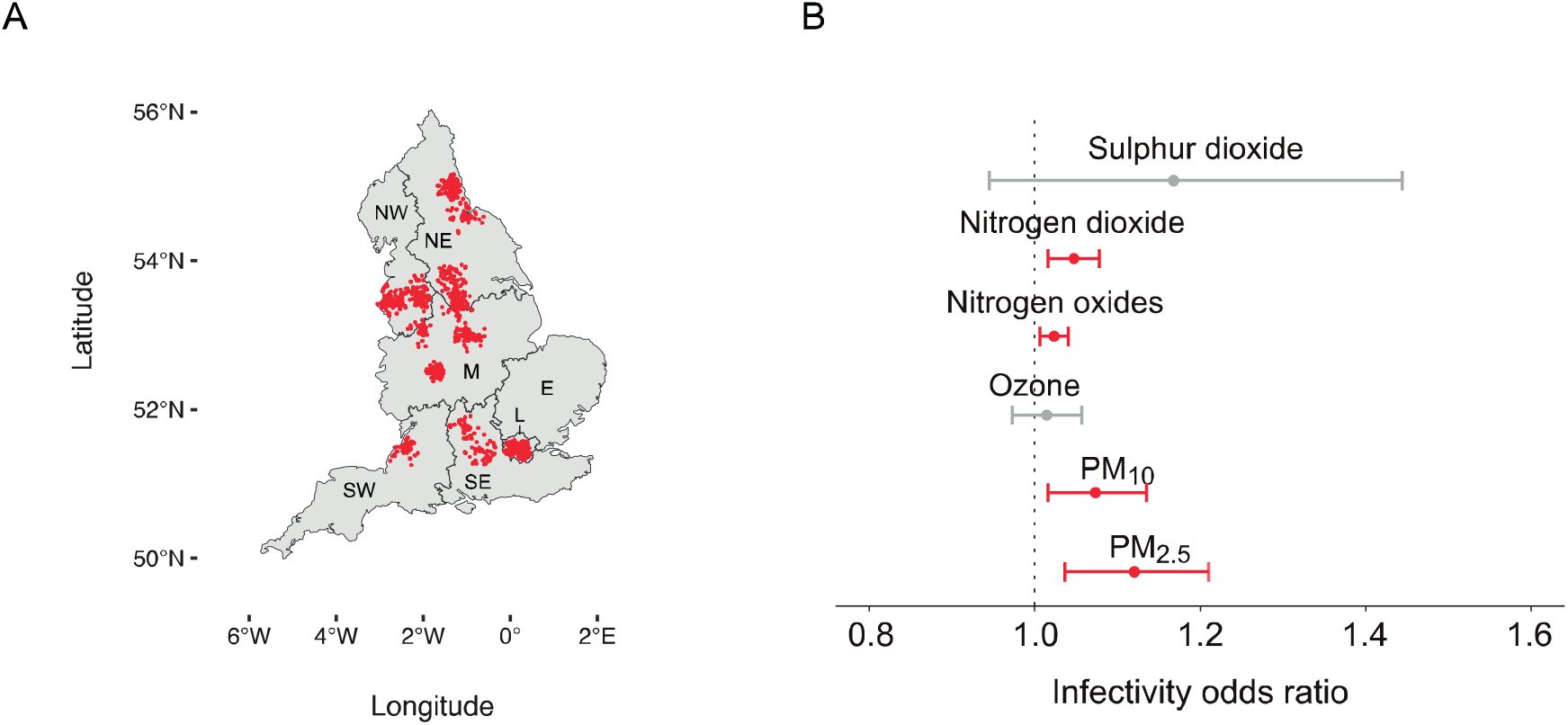
Distribution and infectivity data from the UK Biobank. A) Distribution of UK Biobank subjects included in the current analysis. B) Odds ratios and respective 95% CIs for the relationship between individual exposure to several air pollutants and the number of lab-confirmed COVID-19 cases.

In our model, we accounted for a list of confounding variables (Supplementary Table 1), which we selected based on a previous study ^22^. Our model identified PM_2.5_ and PM_10_ as significant predictors of increased SARS-CoV-2 infectivity (Figure 4B). The odds ratios are 1.120 [CI: 1.036 - 1.211] and 1.074 [CI: 1.017 - 1.136] for PM_2.5_ and PM_10_, respectively. While PM does not predict the numbers of deaths and cases at a subregional level, these pollutants are significant predictors of infectivity at an individual level. Similar to the subregional models (Figure 3), levels of nitrogen oxides and dioxide were predictors of increased infectivity with a lower impact than PMs, with an odds ratio of approximately 1.03 (Figure 4B). Conversely, sulphur dioxide and ozone levels are not significant predictors of infectivity at the individual level, although they are predictors of deaths and cases at the subregional level (Figures 3 and 4B).

We observed an association between current smokers and a lower likelihood of COVID-19 positivity than previous and non-smokers. However, according to our model, population density and predisposing health factors, such as age, sex, diabetes and a previous history of cancer and lung problems, are not predictors of the probability of being infected (Supplementary Table 6).

### Fossil fuel emission levels are linked to pollutants that contribute to increased numbers of COVID-19 deaths

We next collected national emission totals from DEFRA to identify the sources of air pollution associated with COVID-19. While no record for ozone and nitrogen dioxide emissions was identified, road transport accounted for more than 30% of nitrogen oxide emissions from fuel combustion in the UK between 1993 and 2018, while manufacturing and energy industries accounted for approximately 16% and 25%, respectively (Supplementary Table 7). For sulphur dioxide, energy production and transformation emerged as the greatest pollution sources in 2018, accounting for 30% of overall emissions (Supplementary Table 8). We mapped fuel consumption by sector and fuel type in England against air quality values for nitrogen dioxide, nitrogen oxides, sulphur dioxide and ozone to assess individual contributions of fossil fuel consumption sources on air pollution levels. First, we calculated the principal components of each fossil fuel category (road transport, residual fuels and gas consumption) on pollutant levels (Supplementary Table 9). We then employed a Gaussian generalised linear model to characterise the effect of the first two principal components of each category on pollutant levels, after adding population density as a confounding variable. These models indicated that high levels of fossil fuel consumption from on-road vehicles, residual fuels and gas consumption significantly predicted increased nitrogen dioxide, nitrogen oxides, ozone and sulphur dioxide levels (Supplementary Table 9). Based on these results, increases in the levels of each group are associated with increased levels of air pollutants.

We employed an iterative stepwise regression approach that aims to select the most suitable predictors of air pollution to elucidate the effects of individual fossil-fuel burning sources within each category (Supplementary Tables 10-13). We observed significant positive associations between annual average amounts of fuels consumed by A-road buses and nitrogen dioxide, nitrogen oxides and sulphur dioxide (Supplementary Tables 10-13). Contributions from residual fuel types (petroleum, coal, manufactured solid fuels and bioenergy and waste) were disaggregated by sector categories to assess the effects of the industrial, commercial and agricultural sector on air pollution levels. Among residual fuels, petroleum consumed by commercial non-road machinery shows one of the highest positive associations with increased levels of nitrogen oxides (odds ratio = 1.310, 95% CI: 1.092, 1.587), nitrogen dioxide (odds ratio = 1.200, 95% CI: 1.026, 1.414) and ozone (odds ratio = 8.503, 95% CI: 2.029, 35.626), while petroleum consumed by off-road agriculture equipment is negatively associated with the levels of both nitrogen dioxide and nitrogen oxides (Figure 5B-C). In addition, petroleum used for rail transport is a significant contributor to both sulphur dioxide (odds ratio = 1.03, 95% CI: 1.009, 1.051) and nitrogen dioxide (odds ratio = 1.018, 95% CI: 1.000, 1.037) ground-level concentrations (Figure 5A-B), but not ozone levels, where an inverse relationship is identified (Figure 5D). Additionally, domestic consumption of manufactured fossil fuels emerges as one of the strongest predictors of the levels of nitrogen oxides (odds ratio = 1.266, 95% CI: 1.135, 1.416), nitrogen dioxide (odds ratio = 1.210, 95% CI: 1.094, 1.340) and sulphur dioxide (odds ratio = 1.350, 95% CI: 1.212, 1.508), but not ozone levels. Finally, we investigated if weather-corrected levels of domestic and non-domestic gas consumption predict air quality values in England. Potentially toxic ambient concentrations of nitrogen oxides are generated from gas combustion, particularly as a result of indoor household activities ^23^. After accounting for variations in population density, we observed positive associations between domestic gas consumption and both nitrogen oxides and dioxide levels in England (Supplementary Tables 10 and 11).

**Figure 5.**
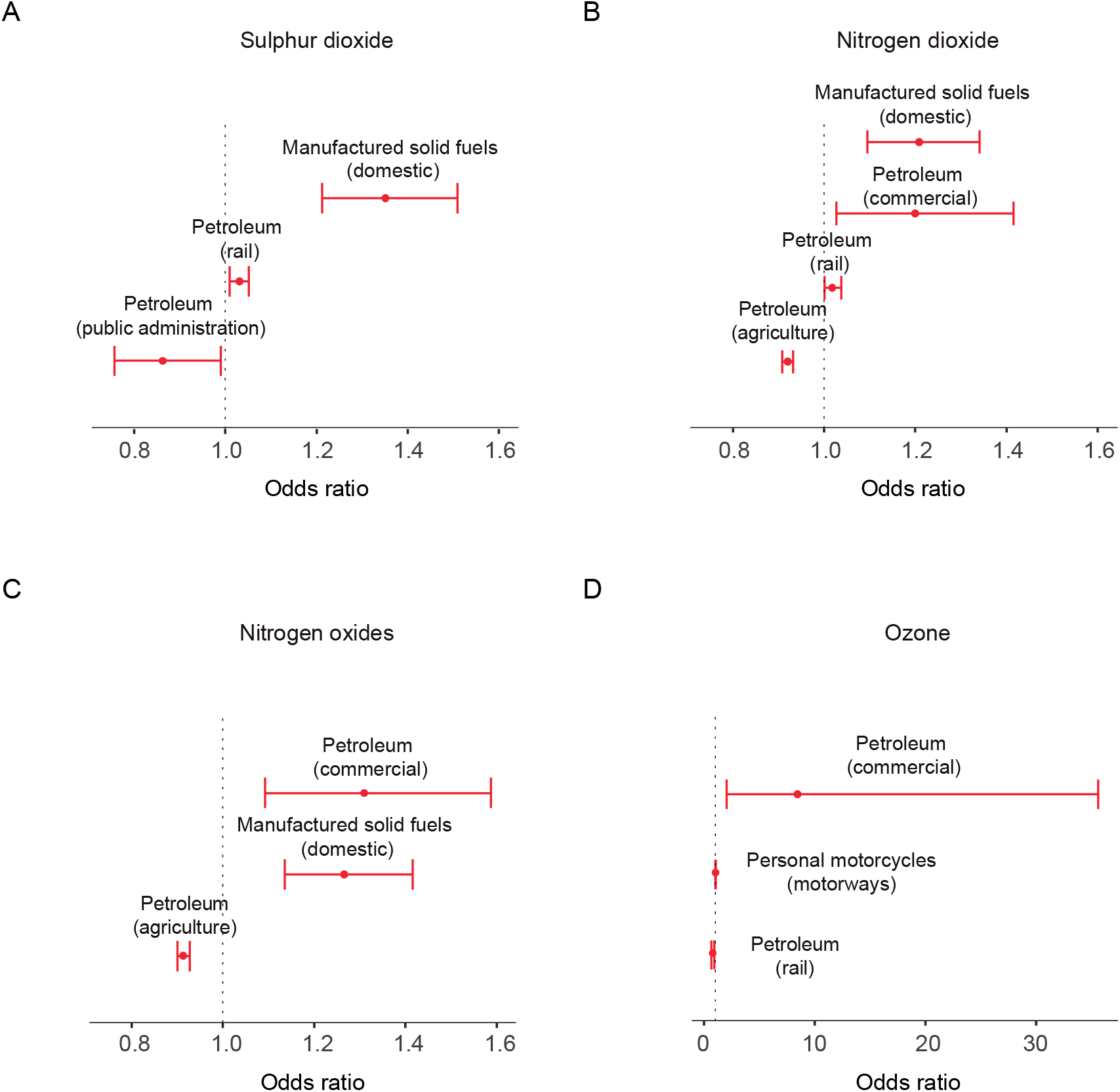
Fossil fuel consumption and air pollution in England. Odds ratios and respective 95% CIs of the effects of fossil fuel consumption stratified by sector and fuel type on A) sulphur dioxide, B) nitrogen dioxide, C) nitrogen oxide and D) ozone levels. Results were normalised to the population density to account for variations in population size across the country. For simplicity, the figure only includes statistically significant (*p* ≤ 0.05) sources of emissions and odds ratios greater than 1.01 or less than 0.99.

## DISCUSSION

Here, we identified associations between air pollution and COVID-19 deaths and cases in England, expanding on previous evidence linking high mortality rates in Europe with increased toxic exposure to air pollutants ^11,24^. Air pollution exposure and health impact estimates have been suggested to mainly depend on the resolution at which they are evaluated ^25^. Therefore, we first calculated the effects of air pollution on COVID-19 mortality and spread using regional, coarse resolution data, and then high-resolution, individual-level observations obtained from the UK Biobank. By employing finer resolution grids, we also show the relative contributions of individual fossil-fuel burning sources to ground-level measurements of air pollutants.

According to our initial findings, regional variations in nitrogen oxide and ozone concentrations in England predict the numbers of COVID-19 cases and deaths, independent of the population density. However, overall uncertainties for modelled exposure estimates at the regional scale ^25^ led us to achieve increased spatial resolution. Using highly granular, local authority-level measurements, we show an association between a 1 μg/m^3^ increase in sulphur dioxide and nitrogen oxide levels with a 17% and approximately 2% increase in COVID-19 mortality, respectively. Notably, these findings are consistent with studies conducted during the previous SARS outbreak, where long-term exposure to air pollutants predicted adverse outcomes in patients infected with SARS in China ^26^. Although nitrogen oxides are key ozone precursors, the relationship between these gases and ozone is nonlinear in ozone chemistry ^27^. Therefore, the negative associations between ozone levels and COVID-19 infection and mortality may be attributed to reduced nitrogen oxide conversion to ozone in urban areas, a phenomenon previously reported for areas with heavy traffic ^20,28^. In addition, given the highly reactive nature of ozone, the inverse relationship between ozone levels and COVID-19 is consistent with increased nitric oxide scavenging close to points of emissions ^29^.

Although the molecular mechanisms underlying the relationship between pollutant exposure and COVID-19 remain to be determined experimentally, they are hypothesised to include the stimulation of chronic, background pulmonary inflammation ^24^. Chamber studies have shown that ambient PM, nitrogen dioxide and sulphur dioxide induce infiltration of the airways by inflammatory cells in healthy volunteers ^30-32^. In addition, exposure to these pollutants may inhibit pulmonary antimicrobial responses, reducing clearance of the virus from the lungs and promoting infectivity. Reduced phagocytic function is well documented after the exposure of macrophages to PM ^1-3^ and is suggested to be the mechanism that enhances viral infection in mice exposed to nitrogen dioxide ^33^.

At the individual level, our UK Biobank model indicated that exposure to PM_2.5_ and PM_10_ increases the risk of COVID-19 infection, in addition to nitrogen oxides, which were previously identified in the regional analysis. This observation conforms to the hypothesis that viruses attach to air pollutants ^34^, potentially explaining the propagation of SARS-CoV-2 and its infectious capacity. Estimations of the viral replication number R_0_ thus must be informed by the local levels of PM. According to our models, demographic features such as age and gender do not alter risk of testing positive for COVID-19. Notably, non-smokers and past smokers are more likely to test positive than current smokers. While we did not investigate the mechanisms by which current smoking protects against hospitalisation due to COVID-19, our findings are consistent with a large body of evidence of a consistently lower prevalence of current smoking among COVID-19 patients and therefore require further investigation ^35^. Although the local authority model suggests a negative association of COVID-19 with PM_2.5_ and PM_10_, the infection model generated using the UK Biobank data produced the opposite results. The conflicting results may arise from diverging testing methodologies in the population samples analysed. While government guidelines in England prioritised testing for symptomatic COVID-19 patients, asymptomatic individuals from the UK Biobank were subjected to COVID-19 testing. Since a large proportion of COVID-19 infections are asymptomatic ^36,37^, the UK Biobank model represents a more accurate estimation of infection.

Based on the purported association between air pollution and COVID-19, we also investigated the contribution of potential sources of key air pollutants to COVID-19 in England. Among the industrial variables, petroleum combustion from non-road commercial machinery emerged as an important predictor of nitrogen dioxide, nitrogen oxide and ozone concentrations. These findings recapitulate previous observations of the estimated concentrations of nitrogen oxides and ozone discharged from stationary combustion sources and off-road mobile machinery ^38,39^. However, operating conditions, engine parameters and vehicle age substantially affect the composition of exhaust emissions ^40^, and thus more detailed information is necessary to construct improved emission models. Among the domestic variables, increased consumption of manufactured solid fuels was associated with higher levels of nitrogen oxides, nitrogen dioxide and sulphur dioxide, indicating a possible link between indoor fuel consumption and air pollution in England. As shown in previous studies, indoor air pollution aggravates the effects of respiratory disorders ^41^, and increasing evidence indicates that home isolation strategies have led to a considerable deterioration of indoor air quality following the COVID-19 outbreak ^42^. As the present study identifies nitrogen oxides and sulphur dioxide as important contributors to COVID-19 mortality, our results are consistent with the hypothesis that indoor air pollution may increase the risk of severe outcomes in COVID-19 patients and thus warrants further attention ^42,43^.

Our findings, supported by results obtained from recent studies conducted in northern Italy ^11^, Europe ^24^, and the USA ^7,44^, suggest that exposure to poor AQ increases the risks of COVID-19 infection and mortality in the UK. Future studies may expand on these observations and address additional confounders, including comorbidities, race, meteorological trends and differences between regional health regulations and their ICU capacities. Nonetheless, air pollution factors should be considered when estimating the SARS-CoV-2 infection rate (R_0_). In addition, our results emphasise the importance of strengthening efforts to tighten air pollution regulations for the protection of human health, both in relation to the COVID-19 pandemic and for the mitigation of potential future diseases.

## Data Availability

Data used for the statistical analyses is available on GitHub

https://github.com/M1gus/AirPollutionCOVID19

https://m1gus.github.io/AirPollutionCOVID19/

## CONFLICTS OF INTEREST

The authors have no conflicts of interest to declare.

## AUTHORS’ CONTRIBUTIONS

MT, YY, RP and LMM planned and designed the study. MT, YY, RP, and NSL collected the data. MT, YY and NSL treated and analysed the data. MT and YY developed the models and wrote the RMD file. MT, YY and RP wrote the manuscript with the support of NSL and LS. LS provided guidance regarding study of air pollution toxicity. MT, YY, RP, LS, NSL and LMM conducted this study while in self-isolation due to the current pandemic.

## ACKNOWLEDGEMENTS

We are grateful to all the staff members with critical functions in administration, operations and logistics at the MRC Toxicology Unit during the present crisis.

## FUNDING

This study is funded by the UK Medical Research Council, intramural project MC_UU_00025/3 (RG94521).

**ABBREVIATIONS**
AIC: Akaike Information Criterion
AQ: Air quality
BEIS: Business, Energy and Industrial Strategy
CI: Confidence intervals
CoV: Coronavirus
COVID-19: Coronavirus disease 19
DEFRA: Department for Environment, Food and Rural Affairs
DfT: Department for Transport
GHGI: Greenhouse Gas Inventory
HGV: Heavy goods vehicle
LGV: Light goods vehicle
MPRN: Meter point reference numbers
NAEI: National Atmospheric Emissions Inventory
NHS: National Health Service
PCA: Principal component analysis
PM: Particulate matter
PM_25_: Particulate matter with an aerodynamic diameter < 2.5 μm
PM_10_: Particulate matter with an aerodynamic diameter < 10.0 μm
PHE: Public Health England
SARS: Severe acute respiratory syndrome
SARS-CoV-2: Severe acute respiratory syndrome coronavirus 2

## Notes

### Competing Interest Statement

The authors have declared no competing interest.

### Funding Statement

This work is funded by the UK Medical Research Council, intramural project MC_UU_00025/3 (RG94521).

